# Replicability and transparency in physical therapy research: Time to wake up

**DOI:** 10.1101/2025.09.23.25336497

**Authors:** François Jabouille, Ata Farajzadeh, Adèle Guillemot Exertier, Jérôme Martine, Natalie Sadek, Leonardo Huet Klinger, Coman Matthew Iliut, Valérie Kapsa, Matthieu P. Boisgontier

**Author notes:** FJ and AF contributed equally to this work.

## Abstract

**Background:** Over the past decade, a replication crisis has been recognized in several scientific disciplines, raising concerns about the validity and reliability of published research and eroding scientific credibility. While this crisis has been widely discussed in other fields, its impact on physical therapy research has received little attention.

**Objective:** This study aimed to provide an overview of the current state of physical therapy research practices that could suggest low replicability.

**Methods:** We assessed the research practices in all original research articles published in official journals of national physical therapy associations in the USA (Physical Therapy), Canada (Physiotherapy Canada), the UK (Physiotherapy), and Australia (Journal of Physiotherapy) between 2022 and 2024.

**Results:** A total of 465 original research articles were identified. Of these, 28% were preregistered, 0% were replication studies or registered reports, and 12% provided open data. Sample size justification was reported in 37% of quantitative studies and 23% of qualitative studies. Despite an average time from submission to publication of 400 days, only 1% of manuscripts were preprinted. Among quantitative studies, 33% stated a hypothesis, while 63% relied on significance testing. Of the reported significant *p*-values, 39% fell within the fragile range (.01≤ p<.05), substantially exceeding the expected 26% under 80% power and suggesting questionable research practices. Hypotheses were supported in 83% of cases, indicating an unreasonably high rate of positive results. A marked difference in the number of *p*-values just above and just below the significance threshold suggested publication bias.

**Conclusion:** These results suggest shortcomings in the reproducibility, replicability, and transparency of the physical therapy literature. Addressing these issues requires the adoption of best practices, including registered reports, justifying sample sizes, publishing negative results, and sharing data and code. Delays in implementing these practices risk exacerbating bias in the literature and undermining the scientific credibility of physical therapy research.

**Statement:** This manuscript is a preprint and is not peer-reviewed. All authors have approved this version of the manuscript.

**Citation:** Jabouille F, Farajzadeh A, Guillemot Exertier A, Martine J, Sadek N, Huet Klinger L, Iliut CM, Kapsa V, & Boisgontier MP. Replicability and transparency in physical therapy research: Time to wake up. *MedRxiv*. 2025.

## INTRODUCTION

In 2011, 270 researchers from 18 countries and 125 institutions began collaborating to replicate 100 studies to test the reliability of prior finding in psychology with different data (Open Science Collaboration, 2015; Nosek et al., 2022). While 97% of the original studies reported statistically significant results, only 36% of the replications did so. Moreover, the effect sizes of the successful replications were often smaller than the original reports (Open Science Collaboration, 2015). Similar replication issues have been reported in other fields, including pharmacology (21%) (Prinz et al., 2011), neuroscience (6%) (Boekel et al., 2015), cancer biology (40%) (Errington et al., 2021), and exercise science (28%) (Murphy et al., 2025).

This poor replicability is often attributed to researcher degrees of freedom (Simmons et al., 2011), which provide scholars with the flexibility to modify hypotheses, study design, data analysis, statistical analysis, and reporting after the results are known (Büttner et al., 2020). If these decisions are not preregistered or made transparent, they create opportunities for questionable research practices, whether deliberate or unintentional, that increase the risk of biased and unreplicable results (John et al., 2012; Boekel et al., 2015; Boisgontier, 2022). Common examples include selectively reporting experiments or variables that produce a desired result (“cherry picking”), presenting unexpected results as if they had been hypothesized a priori (“HARKing”), and conducting various forms of *p*-hacking until statistical significance is achieved (Büttner et al., 2020).

Researchers may engage in these practices for various reasons (Wiggins & Christopherson, 2019). At the system level, these include incentive structures in academia that privilege positive findings (Allen & Mehler, 2019) and the pressures of a “publish-or-perish” culture (Moosa, 2024). At the individual level, contributing factors include cognitive biases, such as the desire to be right (Nickerson, 1998), and the ambition to gain rapid recognition through striking results, even at the expense of methodological rigor (Maltby et al., 2008).

These questionable research practices not only increase the risk of unreplicable results but also contribute to unrealistically high rates of positive results: 88% in applied sciences (Fanelli, 2010), 82% in sports medicine (Büttner et al., 2020), and 81% in exercise sciences (Twomey et al., 2021). Physical therapy is an applied science and shares similar research training practices and academic incentives with sports medicine and exercise sciences. It is therefore urgent to examine whether physical therapy research also exhibit inflated positive result rates, which may signal broader systemic issues in research transparency and methods.

In response to poor replicability, questionable research practices, and unrealistic positive results rates, several fields have recommended the adoption of protocol registration and other open science practices. While these measures have strengthened the literature in psychology (Bogdan, 2025; Bakker et al., 2025), their adoption remains limited in disciplines closely related to physical therapy, such as sports medicine (Bullock et al., 2023), and has not yet been systematically examined in physical therapy research.

The objective of this study was to assess the current state of physical therapy literature and reporting practices by examining key indicators, including the proportion of positive results, replication studies, pre-registrations, registered reports, sample size justification, open data, effect size reporting, fragile *p*-values, *p*-value distribution, and preprinting.

## METHODS

### Sample

A systematic search was conducted to identify the articles reporting original research that were published from 2022 to 2024 in the flagship journals of the leading physical therapy associations in the United States (American Physical Therapy Association), Canada (Canadian Physiotherapy Association), the United Kingdom (Chartered Society of Physiotherapy), and Australia (Australian Physiotherapy Association). These journals provide a sample of the current state of research practices of physiotherapy worldwide. They are, respectively, *PTJ: Physical Therapy & Rehabilitation Journal* (https://academic.oup.com/ptj), *Physiotherapy Canada* (https://www.utpjournals.press/loi/ptc), *Physiotherapy* (https://www.csp.org.uk/professional-clinical/clinical-evidence/physiotherapy-journal), and *Journal of Physiotherapy* (https://www.sciencedirect.com/journal/journal-of-physiotherapy). Only original studies were included in the analyses.

### Data Extraction

Nine coders (FJ, AF, AGE, JM, NS, LHK, CMI, VK) were responsible for data extraction. All coders received standardized training informed by previous research (Twomey et al., 2021, Buttner et al., 2020; Scheel et al., 2021). Each article was independently coded by at least two coders, and any conflicts were resolved by one of the co-first authors (FJ, AF). Data were extracted across seven domains: research topic, data sharing, replication, preregistration, registered report, sample size justification, support for hypotheses, statistical reporting, and preprint.

### Research Topics

All journal articles were categorized into 20 research topics: acute care, balance and falls, cardiovascular and pulmonary, covid, diabetes, education, geriatrics, health policy, implementation science, knowledge translation, measurement, musculoskeletal, neurology, oncology, pain management, pediatrics, prevention and health promotion, professional issues, psychosocial, and women’s health.

### Replication

Replication refers to testing the reliability of a prior finding with different data (Nosek et al., 2022). To identify replication studies, the articles were searched for the string ‘replic^*^‘ (Scheel et al., 2021).

### Preregistration

Preregistration is the practice of making a publicly accessible, time-stamped record of a research plan prior to data collection (Nosek et al., 2018). To assess preregistration, we searched for statements or links indicating that the study had been preregistered.

### Registered Report

A registered report is a publication format with the following submission timeline: Before data collection, authors submit a study protocol outlining their hypotheses, planned methods, and analysis plans. This protocol undergoes peer review. If the protocol is approved, the journal grants in-principle acceptance, committing to publish the final article regardless of whether the hypotheses are supported. Then, authors collect and analyze the data and complete the final report. A second round of peer review then verifies adherence to the registered protocol, ensures that the conclusions are justified, and confirms that the data meet any prespecified quality checks. To assess whether authors could publish registered reports, we examined the websites of the sampled journals to determine whether they offered this type of article.

### Sample Size Justification

To examine sample size justifications, we distinguished between qualitative and quantitative studies, as the two approaches rely on different types of justification. This distinction was made based on an explicit mention of a qualitative methodology in the methods section.

For qualitative studies, sample size justifications were classified into four categories (Vasileiou et al., 2018): (1) data saturation, whereby data collection continues until no new themes emerge; (2) information power, whereby more informative data per participant requires a smaller sample size; (3) data adequacy, reflecting the richness and volume of data; and (4) heuristic, based on generally accepted rules of thumb or norms.

For quantitative studies, justifications were grouped into (1) a priori power analysis, (2) accuracy (i.e., precision of parameter estimates), (3) heuristic, and (4) resource constraints (e.g. limitations due to time, funding, or participant availability) (Lakens, 2022b). For a priori power analyses (1), justifications for the smallest effect size of interest were categorised as benchmark, pilot study, previous study, or meta-analysis (Lakens et al., 2018).

### Hypothesis Statement & Support

To assess the proportion of results that supported the tested hypotheses (i.e., positive results), we first identified articles that stated an explicit hypothesis. If so, we then determined whether the hypothesis was fully supported, partially supported, or not supported, based on the authors’ description of the results. When multiple hypotheses were tested, the primary hypothesis was considered to be the first hypothesis mentioned in the text, unless otherwise clarified (Fanelli, 2010; Scheel et al., 2021).

### Statistical Reporting

To assess the use of null hypothesis significance testing, we searched for related statements and the reporting of *p*-values. We coded whether *p*-values were interpreted as significant and classified their reporting as exact (e.g., p = 0.04), relative (e.g., p < 0.05), or mixed. A report of p < 0.001 was treated as exact, as some statistical software programs do not provide *p*-values below this threshold (Twomey et al., 2021). We also extracted whether effect sizes were reported, including Cohen’s *d*, correlation coefficients, mean differences, and measures of model fit (Twomey et al., 2021).

### Fragile *P*-Values

Questionable research practices could be inferred by the proportion of fragile *p*-values, which refers to a result that could easily change from statistically significant to non-significant with only a small change in the data (Walsh et al., 2014). To extract *p*-values, the unformatted “Results” sections of articles reporting significance tests were extracted, excluding captions, tables, and figures, using code adapted from Bogdan (2025). After *p*-value extraction, the proportion of fragile significant *p*-values was calculated for each article as the number of p-values between.01 and.05 (.01 ≤ p <.05) divided by the number of p values strictly below.05. Only *p*- values reported with equal (=) or less than (< or ≤) signs were included, with ≤ treated as <. For studies that reported all significant *p*-values as “p <.05”, a correction was applied: fragile *p*-value percentages were recomputed based on nearby test statistics when available. If no test statistics were reported, the fragile p-value percentage was set to 51% (Bogdan, 2025).

### *P*-Value Distribution

To assess publication bias, which describes publishing behaviors that give manuscripts that find support for their tested hypotheses a higher chance of being published than manuscripts with “negative” results (Franco et al., 2014; Scheel et al., 2021), we examined the distribution of *p*- values on each side of the significance threshold. First, all extracted exact *p*-values (reported with “=“) were transformed into z-values to facilitate interpretation and plotted to visualize potential differences just above versus just below the significance threshold (Kühberger et al., 2014). Second, relative *p*-values (reported with “<“ or “>“) were categorized into two groups: p <.05 and p >.05.

### Data Sharing

Reproducibility refers to testing the reliability of a prior finding using the same data and the same analysis strategy (Nosek et al., 2022). The most basic requirement for reproducibility is accessible data. To assess data accessibility, we searched for a data availability statement. If a statement was present, we examined whether the dataset was openly accessible, either via a direct link or as a supplementary file.

### Preprints

A preprint is a complete version of a manuscript that authors upload to a public server before peer review (Berg et al., 2016; ASAPbio, 2023). To determine whether an article had been preprinted, we first searched for a corresponding statement. If no such statement was present, we searched Google Scholar using the article title. To estimate the potential for acceleration of knowledge dissemination through preprinting, we calculated the number of days between the journal submission date and the publication of the article online.

## RESULTS

### Types of Articles

Between 2022 and 2024, the four journals published a total of 1,173 articles: 590 in PTJ (American Physical Therapy Association), 168 in Physiotherapy Canada (Canadian Physiotherapy Association), 150 in Physiotherapy (UK Chartered Society of Physiotherapy), and 265 in the Journal of Physiotherapy (Australian Physiotherapy Association). Of these articles, 465 (40%) were original research studies, comprising 391 (84%) quantitative studies and 74 (16%) qualitative studies. The other types of articles included review (14%), perspectives (10%), commentaries (10%), letters to the editor (6%), synopsis (6%), editorials (5%), news (3%), correction (2%), protocols (2%), case reports (2%), guidelines (1%), and historical essays, announcements and association business (<1%) (Figure 1). Among the reviews, 27% were narrative reviews, 72% were systematic reviews, and 43% included a meta-analysis. One study was a meta-analysis of individual patient data without a systematic review.

**Figure 1.**
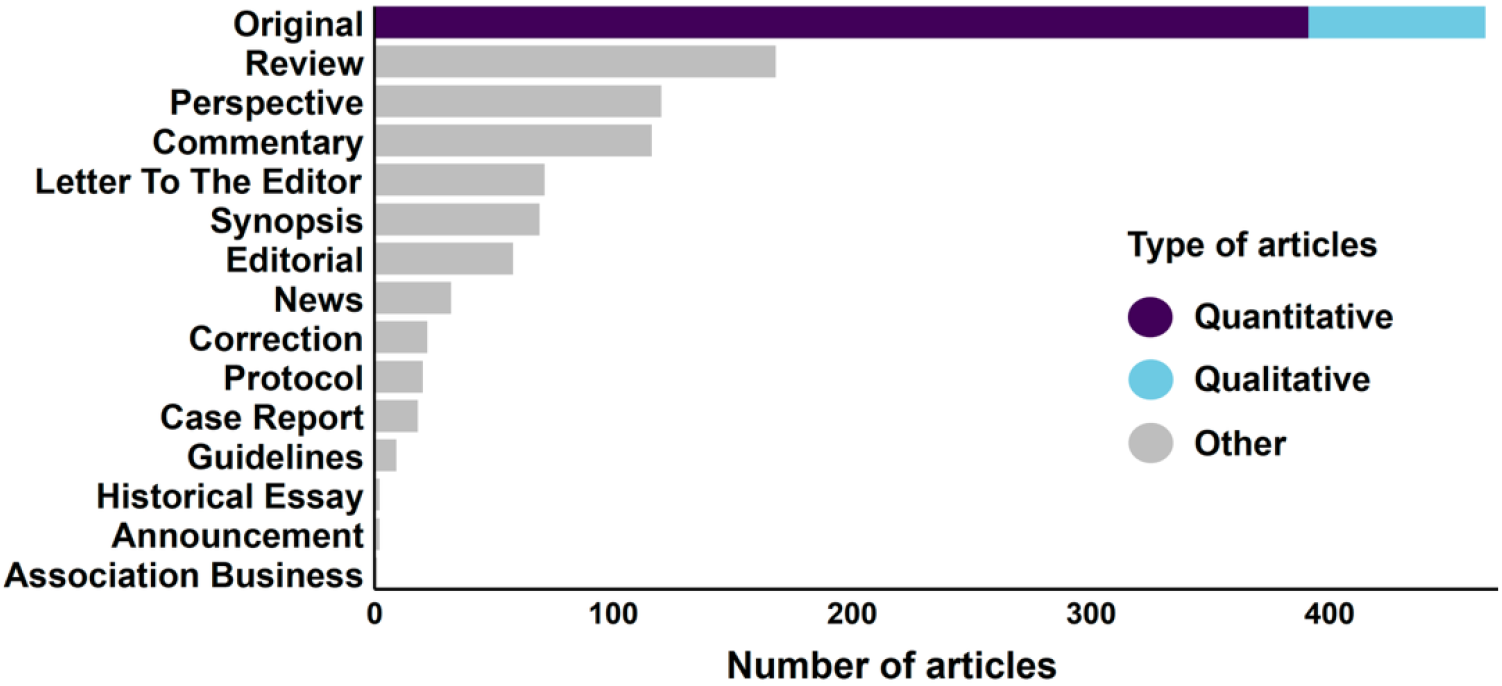
Published articles by type from 2022 to 2024

### Sample Size and Sex Distribution by Research Topic

The median sample size varied considerably by research topic, ranging from 31 participants in the field of neurology to 4,652 in the field of health policy (Table 1). On average, female participants represented 61% (n = 496,294) of the total number of participants in the sampled studies. The proportion of female participants exceeded 70% in studies on diabetes, geriatrics, professional issues, and psychosocial research. In contrast, the proportion of male participants exceeded 55% in cardiovascular and pulmonary research only (Table 1).

**Table 1.** Sample size and sex distribution by research topic.

| Topic | k | n | Median | SD | Min | Max | % Female | % Male |
| --- | --- | --- | --- | --- | --- | --- | --- | --- |
| Acute Care | 8 | 241 | 95 | 136 | 21 | 359 | 46.1 | 53.9 |
| Balance & Falls | 6 | 407 | 34 | 91 | 16 | 253 | 50.1 | 49.9 |
| Cardiovascular & Pulmonary | 22 | 1,116 | 39 | 58 | 8 | 201 | 33.7 | 64.2 |
| COVID | 25 | 129,933 | 75 | 25,180 | 7 | 126,158 | 58.0 | 42.0 |
| Diabetes | 2 | 44 | 40 | 6 | 36 | 44 | 79.5 | 20.5 |
| Education | 41 | 8757 | 53 | 860 | 6 | 5,080 | 67.4 | 31.7 |
| Geriatrics | 14 | 139,326 | 200 | 34,890 | 30 | 131,127 | 74.4 | 25.6 |
| Health Policy | 11 | 337,786 | 4,652 | 196,531 | 32 | 640,901 | 61.2 | 38.8 |
| Implementation Science | 13 | 2,523 | 328 | 1586 | 16 | 5,766 | 59.1 | 40.9 |
| Knowledge Translation | 2 | 156 | 161 | 8 | 156 | 167 | 60.3 | 39.7 |
| Measurement | 39 | 34,748 | 100 | 3,348 | 3 | 20,354 | 57.8 | 41.9 |
| Musculoskeletal | 91 | 97,372 | 84 | 5,797 | 8 | 53,497 | 46.9 | 53.7 |
| Neurology | 57 | 32,075 | 31 | 2,513 | 1 | 14,281 | 51.8 | 48.1 |
| Oncology | 11 | 5,598 | 64 | 1,258 | 17 | 4,264 | 61.9 | 38.7 |
| Pain Management | 30 | 4,201 | 82 | 232 | 7 | 1,143 | 62.9 | 37.6 |
| Pediatrics | 18 | 900 | 37 | 64 | 11 | 245 | 52.3 | 47.7 |
| Prevention & Health Promotion | 9 | 10,784 | 183 | 2,669 | 18 | 8,236 | 51.1 | 48.9 |
| Professional Issues | 34 | 8,237 | 57 | 5,345 | 6 | 26,984 | 72.8 | 25.6 |
| Psychosocial | 7 | 1,029 | 109 | 209 | 10 | 611 | 73.6 | 25.2 |
| Women's Health | 26 | 1,285 | 96 | 875 | 9 | 4,556 | 100.0 | 0.0 |
| **All topics** | **466** | **816,518** | **74** | **33,447** | **1** | **640,901** | **60.7\*** | **39.3\*** |

*Notes: k = number of studies, n = total number of participants, SD = standard deviation, Min = minimum, Max = maximum, \*Studies on women's health are excluded.*

### Replications

None of the 465 original articles were replication studies (Figure 2A).

**Figure 2.**
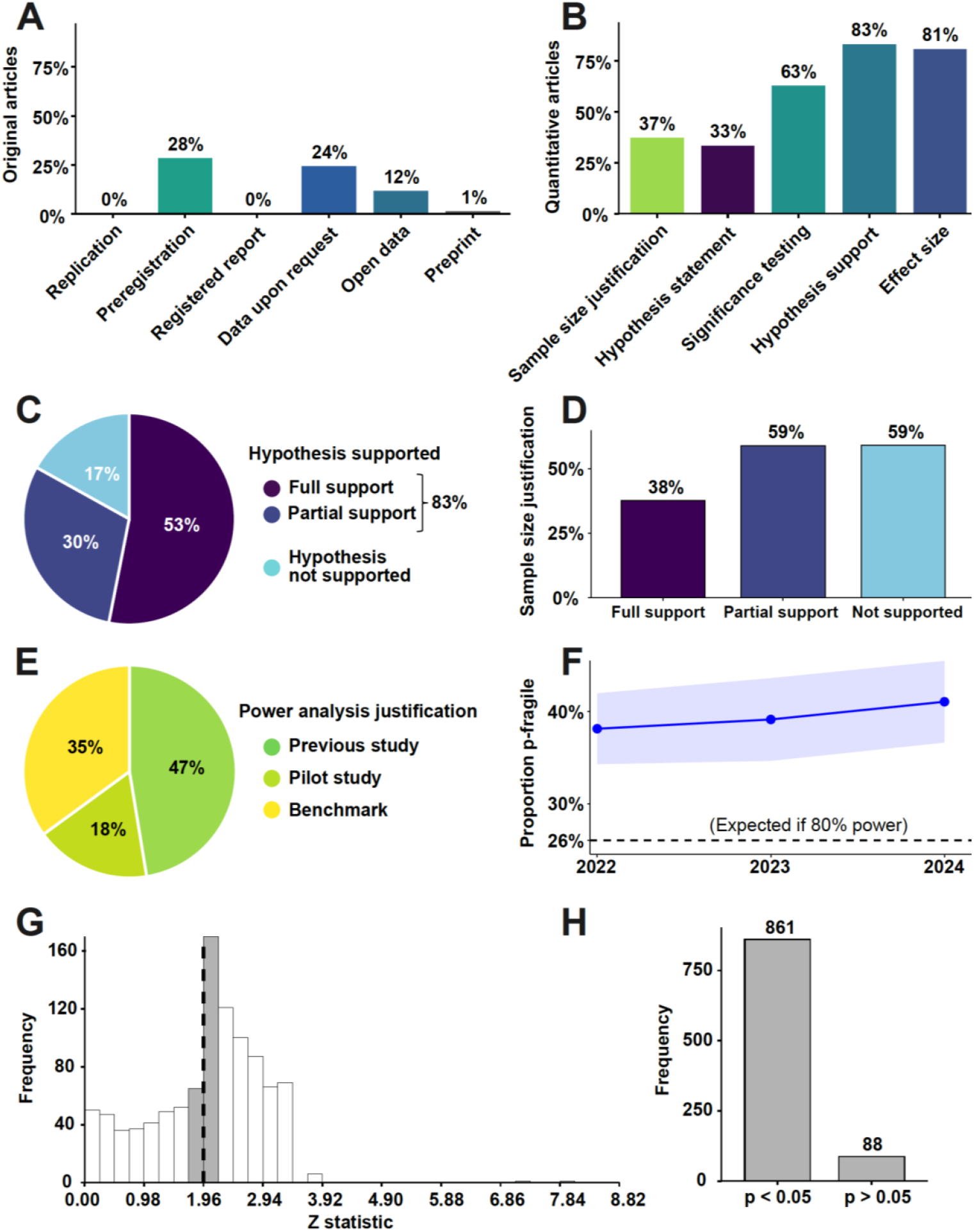
Research practices in physical therapy. (A) Research practices. (B) Quantitative research practices. (C) Types and frequencies of hypothesis support. (D) Sample size justification by type of hypothesis support. (E) Types of power analysis justification. (F) Proportion of fragile *p*-values per article (shaded area represents ± 1 standard error). (G) Distribution of z-transformed exact *p*-value (dashed line represents the z-statistic (1.96) associated with p =.05 significance). (H) Distribution of relative *p*-values.

### Preregistrations

Preregistration statements were present in 28% of the articles (Figure 2A). Registries included the Australian New Zealand Clinical Trials Registry (ANZCTR), Brazilian Registry of Clinical Trial (ReBec), Chinese Clinical Trial Registry (ChiCTR), Clinical trials, Clinical Trials Registry India (CTRI), Netherlands Trial Register (LTR), Open Science Framework OSF), International Clinical Trials Registry Platform (ICTRP), Comet Initiative, International Standard Randomised Controlled Trial Number (ISRCTN), and the Pan-African Clinical Trials Registry (PACTR).

### Registered Reports

None of the sampled journals offered the registered report format for publication. Consequently, the proportion of registered reports was 0% (Figure 2A).

### Sample Size Justification

Sample size justification was reported in 37% of the quantitative articles (Figure 2B) and 23% of the qualitative articles. Among the 37% of quantitative studies justifying their sample size, the most common justification was an a priori power analysis (69%), followed by heuristic (23%), accuracy (6%), and resource constraints (2%). Of those using an a priori power analysis, 47% relied on previous studies to determine the smallest effect size of interest, 35% on benchmark values, 18% on pilot data, and 0% on meta-analyses (Figure 2E). Articles fully supporting their hypothesis justified their sample size in 38% of cases, whereas articles reporting partial or no support both did so 59% of cases (Figure 2D). For qualitative studies, justifications included information power (29%), data saturation (29%), heuristic (24%), and data adequacy (18%).

### Hypothesis Statement and Support

Among the 391 quantitative studies, 130 (33%) explicitly stated at least one hypothesis (Figure 2B). Of these, 83% supported the main hypothesis (Figure 2B): 53% reported full support, 30% partial support, and 17% no support (Figure 2C). Excluding knowledge translation and diabetes due to an insufficient sample size (k = 2), the prevalence of hypothesis testing ranged from 8% in implementation science to 71% in geriatrics (Figure 3A). Hypothesis support varied by topic, ranging from 0% support in psychosocial studies to 100% support in prevention and health promotion, pediatrics, implementation science, education, COVID, and acute care (Figure 3B).

**Figure 3.**
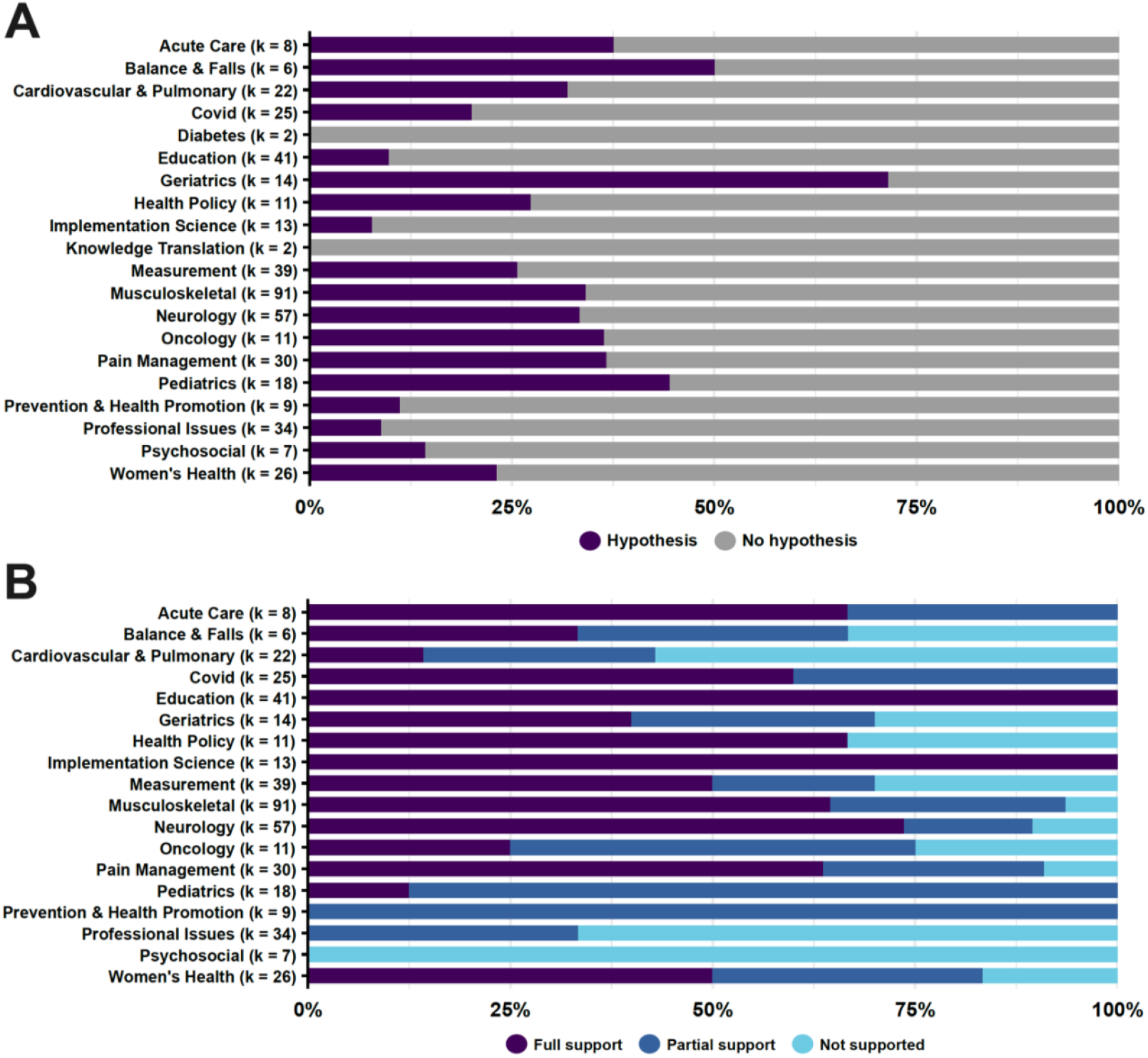
Hypothesis statement (A) and support (B) by research topic.

### Statistical Reporting

Statistical reporting practices were assessed in quantitative original articles only (k = 391). Although 33% of quantitative studies stated a hypothesis, 63% used some form of significance testing (Figure 2B). Notably, 52% of studies without a stated hypothesis still applied significance testing. Among studies using significance testing, 47% reported exact *p*-values, 9% reported relative *p*-values, and 44% reported a combination of both. In addition, 81% of studies reported at least one effect size to contextualize their findings (Figure 2B).

### Fragile *P*-Values

Assuming 80% statistical power, the expected percentage of fragile *p*-values is 26% (Bogdan, 2025). Among the 391 quantitative studies, 204 reported *p*-values in their results section. 39.4% of the significant *p*-values were in the fragile range, with yearly rates of 38.1% in 2022, 39.1% in 2023, and 41.1% in 2024 (Figure 2F). These rates substantially exceed the expected 26%, suggesting that significant findings in the field are more fragile than would be anticipated under adequately powered conditions, which may reflect the influence of questionable research practices.

### Distribution of *P*-Values

The distribution of exact *p*-values showed a drop around the critical z-score of 1.96 (p =.05), with frequencies dropping from 170 in the bin just above to 65 in the bin just below (Figure 2G). The resulting ratio of *p*-values just reaching significance to those just failing is 2.6:1, an outcome that is a highly unlikely under normal conditions. Further, among relative *p*-values, 861 fell in the p <.05 category and 88 in the p >.05 category (Figure 2H). Together, these results suggest the presence of publication bias.

### Data Sharing

Although 36% of the articles included a data accessibility statement, only 12% made their data openly available (Figure 2A). The remaining 24% were “available upon reasonable request”.

### Preprints & Time to Publication

Six manuscripts (1.3%) were available as preprints (Figure 2A). The average time from submission of a manuscript to publication in a journal was 400 ± 199 days.

## DISCUSSION

### Main Findings

To assess the current state of research and reporting practices, we systematically evaluated 465 original articles published between 2022 and 2024 in the flagship journals of the leading physical therapy associations in the United States, Canada, the United Kingdom, and Australia. We observed that 83% of the studies confirmed their main hypothesis, which is a highly unlikely positive result rate. Only 37% of the quantitative studies and 23% of the qualitative studies justified their sample size, which increases the risk of low-powered studies. No registered reports or replication studies were conducted, and pre-registration was low (28%), paving the way for post-hoc questionable research practices. Data were openly available in only 12% of articles, which limits reproducibility. The mean proportion of fragile *p*-values per article was 39%, exceeding the expected percentage of 26% and suggesting questionable research practices. An inordinately high number of p-values just passed the boundary of significance, suggesting publication bias. Only six articles were preprinted, delaying the dissemination of research results.

### Unrealistically High Rate of Hypothesis Support

The positive result rate of 83% that we identified in the physical therapy literature was comparable to the rates reported in sports medicine (82%) (Büttner et al., 2020), exercise sciences (81%) (Twomey et al., 2021), and applied sciences (88%) (Fanelli, 2010). This extremely high rate of hypothesis support may be partly explained by questionable research practices, which have been consistently admitted by scientists in several fields (John et al., 2012; Fiedler & Schwarz, 2016; Agnoli et al., 2017; Fraser et al., 2018; Makel et al., 2021; Gopalakrishna et al., 2022; Isbell et al., 2022; Schneider et al., 2024). Another possible explanation is publication bias, which can be indirectly evidenced by the analysing *p*-value distributions (Kühberger et al., 2014; Borg et al., 2023). In our study, these distributions clearly suggested the presence of publication bias. To reduce this bias and the incentive to publish positive results only, physical therapy journals could explicitly state that they accept negative results and dedicate a section to these studies, similar to what other journals have implemented (PLoS One, 2025).

### Limited Use of Registration Practices

Another way to reduce this publication bias and to reduce the possibility for questionable research practices is to encourage authors to register their research plans prior to data collection through preregistration (Kaplan & Irvin, 2015; Zarin et al., 2019) or registered reports (Allen & Mehler, 2019; Scheel et al., 2021). These practices aim to prevent the methods from being influenced by the observed data (Lakens et al., 2024). The value of registered reports is clearly demonstrated in psychology, where 96% of standard reports yielded positive results (Scheel et al., 2021). In contrast, when the nature of the results became irrelevant to acceptance for publication through the registered report format, the proportion of positive results dropped to 44%, which more closely reflects the proportion of true effects one would expect in reality (Scheel et al., 2021).

None of the journals of the leading physical therapy associations we included in this study offered researchers the option of submitting their work as a registered report, and the rate of preregistration was slightly above one quarter of the articles (28%). This rate is similar to those reported for clinical trials (26%) (Bramley et al., 2025) and higher than in the fields of exercise sciences (9%) (Twomey et al., 2021) or psychology (3%) (Hardwicke et al., 2022). To reduce publication bias and the opportunities for questionable research practices, journals should offer registered reports as an article type, enabling authors to obtain in-principle acceptance based on their peer-reviewed protocol. Journals could also partner with platforms that conduct independent peer reviews of registered reports (e.g., Peer Community In Registered Reports: https://rr.peercommunityin.org/PCIRegisteredReports/about/about).

### Replicability: A Blind Spot and a Crisis to Come

The high rate of positive findings reported here calls into question the replicability of physical therapy research. Yet, this replicability is not assessed. We found no replication studies in the sampled journals over a 3-year period. However, replicability can be studied indirectly by examining the distribution of *p*-values (Bogdan, 2025). For example, studies with 80% statistical power should show fewer than 26% of *p*-values within the.01 ≤ p ≤.05 interval (Bogdan, 2025). Here, on average, 39% of the p-values per article were fragile. This percentage is higher than that experienced by psychology more than a decade ago (32%) between 2004 and 2011, before their replicability crisis (Bogdan, 2025). Since then, the robustness of psychology research has gradually increased, with the percentage of fragile *p*-values nearly reaching 26% (Bogdan, 2025). Our results suggest that the proportion of replicable studies in physical therapy may be comparable to or lower than the 36% rate observed in psychology before its replicability crisis (Open Science Collaboration, 2015).

### Risk of False Findings

Sample size is critical for reproducibility because it directly affects the risk of obtaining false results (Brysbaert, 2019). Justifying the sample size enables readers to evaluate this risk and determine whether it is acceptable given the knowledge contributed by the study. However, in the four journals we analyzed, only 37% of articles provided a justification for their sample size. Consequently, in approximately two thirds of the articles, readers cannot determine whether the sample size was adequate to reliably detect the tested effects, nor whether the decision not to assess this risk was reasonable. Moreover, the accuracy of a power analysis depends on the reliability of the effect size used. Yet, none of the studies relied on the strongest form of justification (i.e., meta-analytic estimates), and 35% relied on conventional benchmarks rather than empirical evidence, an approach that is widely discouraged as it offers no study-specific rationale (Lakens, 2022b). To address this issue, physical therapy journals should explicitly encourage authors to justify their sample size and to provide sufficient information to enable their a priori power analyses to be reproduced (Mckay et al., 2023).

### Low Reproducibility

Only 12% of articles made their data directly available in a public repository or as supplementary material, meaning that reproducibility could be directly assessed in just over one-tenth of the physical therapy literature. An additional 24% of articles included a data availability statement indicating that datasets were available upon reasonable request. However, such statements have proven largely ineffective: in a study of 3,556 BioMed Central articles, 93% of authors who made these statements either did not respond or declined to share their data (Gabelica et al., 2022). To improve reproducibility in physical therapy research, journals could adopt a default open data policy, permitting exceptions only when authors provide documented evidence that data sharing is restricted by an ethics boards or third-party agreements. Evidence from the social sciences suggests that such policies would be well received, with 88% of authors expressing support for the posting of data or code online (Ferguson et al., 2023). Moreover, established journals such as the *British Medical Journal* already require data and code to be shared (Loder et al., 2024).

Another barrier to reproducibility is that 53% of articles reported relative or mixed *p*-values, which limits the feasibility of meta-analyses and secondary analyses. Following the lead of several established medical journals (Aguinis et al., 2021), physical therapy journals should require reporting of exact *p*-values in accordance with standard conventions for decimal formatting (Lang & Altman, 2015).

### Disconnect Between Hypothesis Statement & Statistical Reporting

Only 33% of the quantitative articles explicitly stated a hypothesis, which is substantially lower than in related fields, such as exercise sciences (64%; Twomey et al., 2021) and sports medicine (60%; Büttner et al., 2020). Together with the relatively high prevalence of effect size reporting (81%), this difference may reflect recent changes in the statistical reporting policies of physical therapy journals. In 2022, an editorial published in several journals, including three of the four journals in our sample, informed researchers that some physical therapy journals in the International Society of Physiotherapy Journal Editors (ISPJE) would expect manuscripts to use estimation methods instead of null hypothesis significance testing (Elkins et al., 2022). This recommendation was met with some critique (Lakens, 2022a; Lohse, 2022; Tenan & Caldwell, 2022) and has since evolved into more nuanced guidelines, emphasizing that “*p-*values are only one piece of information that needs to be considered jointly with the uncertainty of estimates, the study context, the costs of different types of errors, and the prior probability of the hypothesis itself” (Lohse & Kliethermes, 2025). These guidelines align with the authors’ apparent reluctance to abandon null hypothesis significance testing altogether, as suggested by its continued use in over half of studies that did not explicitly state a hypothesis.

### Slow Knowledge Dissemination

Preprints serve as a public record of the author’s original manuscript (Butler & Boisgontier, 2025), contributing to transparency by making modifications introduced during peer review and editorial processes visible, and potentially reducing bias at these stages. By bypassing peer-review and publication delays, preprints enable the early dissemination of research results, accelerating scientific discovery (Maggio et al., 2018). Modeling the effects of removing these delays over a 10-year period suggests that discovery could be accelerated up to fivefold (Quake, 2017). These benefits have led several national funding agencies to recognize and encourage preprinting, including those in the countries of the sampled journals: the United States (National Institutes of Health, 2025), Canada (Canadian Institutes of Health Research, 2023), the United Kingdom (Wellcome, 2025), and Australia (Australian Research Council, 2021). Despite this, our results indicate that only 1.3% of physical therapy research is preprinted, delaying knowledge dissemination by over a year, given that the average time from submission to journal publication is 400 days. To promote preprinting, journals should not only allow the submission of preprinted manuscript, but also explicitly encourage this practice in their author guidelines. Additionally, journals could facilitate submissions by enabling direct transfers from preprint repositories, as the Physical Therapy & Rehabilitation Journal (PTJ) does with MedRxiv (https://www.medrxiv.org/submit-a-manuscript).

### Limitation and Future Considerations

This study has several limitations that should be noted. First, the study is based on four journals. Although they are the official journal of the main physical therapy associations in their respective countries, future studies should expand the breadth of article sources. Second, we assessed journals that are published within developed countries, which limits the generalizability of the findings. Future research should examine research practices in physical therapy within a global context. Finally, we assessed research practices indirectly, based on articles. Future research should investigate these practices more directly by surveying researchers in physical therapy.

### Conclusions

Our results suggest that the replicability and reproducibility of physical therapy research is deficient. To address these issues, we recommend that the field urgently consider adopting registered reports, mandating open data and code sharing, encouraging preprints, and providing clear author guidance on statistical reporting. Implementing these measures could improve transparency, reduce publication bias, and strengthen the overall credibility of physical therapy research.

## Data Availability

Data, code, and material used for this study are publicly available: https://doi.org/10.5281/zenodo.17138184

## ARTICLE INFORMATION

### Authorship Contribution Statement

Based on the Contributor Roles Taxonomy (CRediT) (Allen et al., 2019), individual author contributions to this work are as follows: François Jabouille, Ata Farajzadeh: Investigation, Formal Analysis, Visualization, Writing – Original Draft; Writing – Review and Editing; Adèle Guillemot Exertier, Jérôme Martine, Natalie Sadek, Leonardo Huet Klinger, Matthew Coman Iliut, Valérie

Kapsa: Investigation; Matthieu Boisgontier: Conceptualization, Methodology, Investigation, Data Curation, Formal Analysis, Visualization, Writing – Original Draft, Writing – Review and Editing, Supervision, Project Administration, Funding Acquisition.

## REFERENCES

Agnoli, F., Wicherts, J. M., Veldkamp, C. L. S., Albiero, P., & Cubelli, R. (2017). Questionable research practices among Italian research psychologists. PLoS One, 12(3), e0172792. 10.1371/journal.pone.0172792

Aguinis, H., Vassar, M., & Wayant, C. (2021). On reporting and interpreting statistical significance and p values in medical research. BMJ Evidence-Based Medicine, 26(2), 39–42. 10.1136/bmjebm-2019-111264

Allen, C., & Mehler, D. M. A. (2019). Open science challenges, benefits and tips in early career and beyond. PLoS Biology, 17(5), e3000246. 10.1371/journal.pbio.3000246

ASAPbio. (2023). Preprints. https://asapbio.org/preprint-info (accessed August 2025)

Australian Research Council. (2021). Open Access Policy (Version 2021.1). Policy and Strategy Branch. https://www.arc.gov.au/sites/default/files/2022-06/Open%20Access%20Policy%20Version%202021.1.docx

Bakker, M., Marszalek, J. M., & Lê, T. T. (2025). Increasing sample sizes in psychology over time. PsyArxiv. 10.31234/osf.io/vwub3_v1

Berg, J. M., Bhalla, N., Bourne, P. E., et al. (2016). Preprints for the life sciences. Science, 352, 899–901. 10.1126/science.aaf9133

Boekel, W., Wagenmakers, E.-J., Belay, L., et al. (2015). A purely confirmatory replication study of structural brain-behavior correlations. Cortex, 66, 115–133. 10.1016/j.cortex.2014.11.019

Bogdan, P. C. (2025). One decade into the replication crisis, how have psychological results changed? Advances in Methods and Practices in Psychological Science, 8(2), 1–14. 10.1177/25152459251323480

Boisgontier, M. P. (2022). Research integrity requires to be aware of good and questionable research practices. European Rehabilitation Journal, 2(1), 1–3. 10.52057/erj.v2i1.24

Borg, D. N., Barnett, A. G., Caldwell, A. R., White, N. M., & Stewart, I. B. (2023). The bias for statistical significance in sport and exercise medicine. Journal of Science and Medicine in Sport, 26(3), 164–168. 10.1016/j.jsams.2023.03.002

Bramley, P., Bird, C., Badgett, R., & DeVito, N. J. (2025). Patterns of preregistration and publication of trials in Cochrane systematic reviews of interventions. Journal of Clinical Epidemiology, 187, 111958. 10.1016/j.jclinepi.2025.111958

Brysbaert, M. (2019). How many participants do we have to include in properly powered experiments? A tutorial of power analysis with reference tables. Journal of Cognition, 2(1), 1–16. 10.5334/joc.72

Bullock, G. S., Ward, P., Impellizzeri, F. M., Kluzek, S., Hughes, T., Hillman, C., …, Collins, G.S. (2023). Up front and open? Shrouded in secrecy? Or somewhere in between? A meta-research systematic review of open science practices in sport medicine research. Journal of Orthopaedic & Sports Physical Therapy, 53(12), 735–747. 10.2519/jospt.2023.12016

Butler, L. A., & Boisgontier, M. P. (2025). Rethinking where and how we publish in health sciences. SportRxiv. 10.51224/SRXIV.600

Büttner, F., Toomey, E., McClean, S., Roe, M., & Delahunt, E. (2020). Are questionable research practices facilitating new discoveries in sport and exercise medicine? The proportion of supported hypotheses is implausibly high. British Journal of Sports Medicine, 54(22), 1365–1371. 10.1136/bjsports-2019-101863

Canadian Institutes of Health Research. (2023). Preprints at CIHR. https://www.irsc.gc.ca/e/50574.html

Elkins, M. R., Pinto, R. Z., Verhagen, A., Grygorowicz, M., Söderlund, A., Guemann, M., … Sheikh, U. (2022). Statistical inference through estimation: recommendations from the International Society of Physiotherapy Journal Editors. Physical Therapy, 102(6). 10.1093/ptj/pzac066

Errington, T. M., Mathur, M., Soderberg, C. K., Denis, A., Perfito, N., Iorns, E., & Nosek, B. A. (2021). Investigating the replicability of preclinical cancer biology. eLife, 10, e71601. 10.7554/eLife.71601

Fanelli, D. (2010). “Positive” results increase down the hierarchy of the sciences. PLoS One, 5(4), e10068. 10.1371/journal.pone.0010068

Ferguson, J., Littman, R., Christensen, G., Paluck, E. L., Swanson, N., Wang, Z., …, & Pezzuto, J. H. (2023). Survey of open science practices and attitudes in the social sciences. Nature Communications, 14(1), 5401. 10.1038/s41467-023-41111-1

Fiedler, K., & Schwarz, N. (2016). Questionable research practices revisited. Social Psychological and Personality Science, 7(1), 45–52. 10.1177/1948550615612150

Fraser, H., Parker, T., Nakagawa, S., Barnett, A., & Fidler, F. (2018). Questionable research practices in ecology and evolution. PLoS One, 13(7), e0200303. 10.1371/journal.pone.0200303

Gabelica, M., Bojčić, R., & Puljak, L. (2022). Many researchers were not compliant with their published data sharing statement: a mixed-methods study. Journal of Clinical Epidemiology, 150, 33–41. 10.1016/j.jclinepi.2022.05.019

Gopalakrishna, G., Ter Riet, G., Vink, G., Stoop, I., Wicherts, J. M., & Bouter, L. M. (2022). Prevalence of questionable research practices, research misconduct and their potential explanatory factors: a survey among academic researchers in The Netherlands. PLoS One, 17(2), e0263023. 10.1371/journal.pone.0263023

Hardwicke, T. E., Thibault, R. T., Kosie, J. E., Wallach, J. D., Kidwell, M. C., & Ioannidis, J. P. A. (2022). Estimating the prevalence of transparency and reproducibility-related research practices in psychology (2014-2017). Perspectives on Psychological Science, 17(1), 239–251. 10.1177/1745691620979806

Isbell, D.R., Brown, D., Chen, M., Derrick, D.J., Ghanem, R., Arvizu, M.N.G., & Plonsky, L. (2022). Misconduct and questionable research practices: the ethics of quantitative data handling and reporting in applied linguistics. Modern Language Journal, 106, 172–195. 10.1111/modl.12760

John, L. K., Loewenstein, G., & Prelec, D. (2012). Measuring the prevalence of questionable research practices with incentives for truth telling. Psychological Science, 23(5), 524–532. 10.1177/0956797611430953

Kaplan, R. M., & Irvin, V. L. (2015). Likelihood of null effects of large NHLBI clinical trials has increased over time. PLoS One, 10(8), e0132382. 10.1371/journal.pone.0132382

Kühberger, A., Fritz, A., & Scherndl, T. (2014). Publication bias in psychology: a diagnosis based on the correlation between effect size and sample size. PloS one, 9(9), e105825. 10.1371/journal.pone.0105825

Lang, T. A., & Altman, D. G. (2015). Basic statistical reporting for articles published in biomedical journals: the “Statistical Analyses and Methods in the Published Literature” or the SAMPL Guidelines. International Journal of Nursing Studies, 52(1), 5–9. 10.1016/j.ijnurstu.2014.09.006

Lakens, D. (2022a). Correspondence: reward, but do not yet require, interval hypothesis tests. Journal of Physiotherapy, 68(3), 213–214. 10.1016/j.jphys.2022.06.004

Lakens, D. (2022b). Sample size justification. Collabra Psychology. 8(1), 33267. 10.1525/collabra.33267

Lakens, D., Scheel, A. M., & Isager, P. M. (2018). Equivalence testing for psychological research: a tutorial. Advances in Methods and Practices in Psychological Science, 1(2), 259–269. 10.1177/2515245918770963

Lakens, D., Mesquida, C., Rasti, S., & Ditroilo, M. (2024). The benefits of preregistration and Registered Reports. Evidence-Based Toxicology, 2(1), 2376046. 10.1080/2833373X.2024.2376046

Loder, E., Macdonald, H., Bloom, T., & Abbasi, K. (2024). Mandatory data and code sharing for research published by The BMJ. British Medical Journal, 384, q324. 10.1136/bmj.q324

Lohse, K. (2022). No estimation without inference: a response to the International Society of Physiotherapy Journal Editors. Communications in Kinesiology, 1(4), 1–10. 10.51224/SRXIV.178

Lohse, K. R., & Kliethermes, S. (2025). Approaching Significance: Statistical Guidance for Authors and Reviewers. Journal of Neurologic Physical Therapy: JNPT. 10.1097/NPT.0000000000000526

Maggio, L. A., Artino Jr, A. R., & Driessen, E. W. (2018). Preprints: facilitating early discovery, access, and feedback. Perspectives on Medical Education, 7(5), 287–289. 10.1007/s40037-018-0451-8

Makel, M. C., Hodges, J., Cook, B. G., & Plucker, J. (2021). Both questionable and open research practices are prevalent in education research. Educational Researcher, 50(8), 493–504. 10.3102/0013189X211001356

Maltby, J., Day, L., Giles, D., Gillett, R., Quick, M., Langcaster-James, H., & Linley, P. A. (2008). Implicit theories of a desire for fame. British Journal of Psychology, 99(2), 279–292. 10.1348/000712607X224661

McKay, B., Bacelar, M. F., & Carter, M. J. (2023). On the reproducibility of power analyses in motor behavior research. Journal of Motor Learning and Development, 11(1), 29–44. 10.1123/jmld.2022-0061

Moosa, I. A. (2024). Publish or perish: Perceived benefits versus unintended consequences. Edward Elgar Publishing.

Murphy, J., Caldwell, A. R., Mesquida, C., Ladell, A. J., Encarnación-Martínez, A., Tual, A., …, Warne, J.P. (2025). Estimating the replicability of sports and exercise science research. Sports Medicine. 10.1007/s40279-025-02201-w

Nickerson, R. S. (1998). Confirmation bias: a ubiquitous phenomenon in many guises. Review of General Psychology, 2(2), 175–220. 10.1037/1089-2680.2.2.175

National Institutes of Health. (2025). NIH preprint pilot. PubMed Central. https://pmc.ncbi.nlm.nih.gov/about/nihpreprints/

Nosek, B. A., Ebersole, C. R., DeHaven, A. C., & Mellor, D. T. (2018). The preregistration revolution. Proceedings of the National Academy of Sciences, 115(11), 2600–2606. 10.1073/pnas.1708274114

Nosek, B. A., Hardwicke, T. E., Moshontz, H., Allard, A., Corker, K. S., Dreber, A., & Vazire, S. (2022). Replicability, robustness, and reproducibility in psychological science. Annual Review of Psychology, 73, 719–748. 10.1146/annurev-psych-020821-114157

Open Science Collaboration. (2015). Estimating the reproducibility of psychological science. Science, 349(6251), aac4716. 10.1126/science.aac4716

PLoS One. Criteria for publication. Retrieved 19 September 2025, from https://journals.plos.org/plosone/s/criteria-for-publication

Prinz, F., Schlange, T., & Asadullah, K. (2011). Believe it or not: How much can we rely on published data on potential drug targets? Nature Reviews Drug Discovery, 10(9), 712. 10.1038/nrd3439-c1

Quake, S. (2017). Stanford Medicine Big Data: Precision Health 2017. [Video]. YouTube. https://www.youtube.com/watch?v=zt9hlbet2Lk&t=402s

Scheel, A. M., Schijen, M. R. M. J., & Lakens, D. (2021). An excess of positive results: comparing the standard psychology literature with registered reports. Advances in Methods and Practices in Psychological Science, 4(2), 1–12. 10.1177/25152459211007467

Schneider, J. W., Allum, N., Andersen, J. P., Petersen, M. B., Madsen, E. B., Mejlgaard, N., & Zachariae, R. (2024). Is something rotten in the state of Denmark? Cross-national evidence for widespread involvement but not systematic use of questionable research practices across all fields of research. PLoS One, 19(8), e0304342. 10.1371/journal.pone.0304342

Simmons, J. P., Nelson, L. D., & Simonsohn, U. (2011). False-Positive psychology: undisclosed flexibility in data collection and analysis allows presenting anything as significant. Psychological Science, 22(11), 1359–1366. 10.1177/0956797611417632

Tenan, M., & Caldwell, A. (2022). Confidence intervals and smallest worthwhile change are not a panacea: a response to the International Society of Physiotherapy Journal Editors. Communications in Kinesiology, 1(4), 1–10. 10.51224/cik.2022.45

Twomey, R., Yingling, V., Warne, J., Schneider, C., McCrum, C., Atkins, W., …, & Caldwell, A. (2021). The nature of our literature: a registered report on the positive result rate and reporting practices in kinesiology. Communications in Kinesiology, 1(3), 1–17. 10.51224/cik.v1i3.43

Vasileiou, K., Barnett, J., Thorpe, S., & Young, T. (2018). Characterising and justifying sample size sufficiency in interview-based studies: systematic analysis of qualitative health research over a 15-year period. BMC Medical Research Methodology, 18(1), 148. 10.1186/s12874-018-0594-7

Walsh, M., Srinathan, S. K., McAuley, D. F., Mrkobrada, M., Levine, O., Ribic, C., …, & Devereaux, P. J. (2014). The statistical significance of randomized controlled trial results is frequently fragile: a case for a Fragility Index. Journal of Clinical Epidemiology, 67(6), 622–628. 10.1016/j.jclinepi.2013.10.019

Wellcome. (2025). Open access policy. https://wellcome.org/research-funding/guidance/ending-a-grant/open-access-guidance/open-access-policy#preprints-24fc

Wiggins, B. J., & Christopherson, C. D. (2019). The replication crisis in psychology: an overview for theoretical and philosophical psychology. Journal of Theoretical and Philosophical Psychology, 39(4), 202–217. 10.1037/teo0000137

Zarin, D. A., Fain, K. M., Dobbins, H. D., Tse, T., & Williams, R. J. (2019). 10-year update on study results submitted to ClinicalTrials.gov. New England Journal of Medicine, 381(20), 1966–1974. 10.1056/NEJMsr1907644

